# Development and validation of a digital twin for the analog scoliometer

**DOI:** 10.1101/2023.11.30.23298978

**Authors:** Sinduja Suresh, Annabelle Stubbs, Amir Amiri, Maree T Izatt, Robert D Labrom, Geoffrey N Askin, J Paige Little

## Abstract

Scoliosis is a complex 3D spine deformity characterised by an abnormal lateral curvature of the spine and associated rotation of the spine and ribcage. The rotational aspect of scoliosis is most commonly quantified in the Adam’s forward flexed position using an analog scoliometer. The scoliometer has a known user error of 5-8°, which is largely dependent on examiner experience, location of curve, patient positioning and BMI. The device is also limited by the 30° scale and parallax errors. Additionally, the scoliometer loses accuracy when the patient’s torso cannot be positioned parallel to the ground . This study describes the development of the first digital twin for the analog scoliometer to enable fast, gravity-independent reliable and accurate digital measurements of the Angle of Torso Rotation (ATR) from patient-specific 3D virtual models.

A robust semi-automated algorithm of generative design which measures ATR from surface topography was developed. With an operating time of just a few seconds, it provides quick and reliable ATR measurements from simple parametric user inputs. 150 calibrated 3D virtual models of AIS patients treated at the Queensland Children’s Hospital Spine Clinic (QCHSC) obtained from our existing database of 3D surface scans (3DSS) and healthy non-scoliotic controls recruited for this study were used to validate the digital scoliometer tool.

The tool showed excellent reliability in both intra-user (0.99) and inter-user (0.98) conditions. The digital values had a high positive correlation (0.897) and agreement (92.7%) with the analog ATR measurements made clinically. The tool also showed high sensitivity (95.83%) and specificity (76.76%). The development and validation of this virtual digital tool is significant for telehealth implementation in paediatric spine deformity management and is expected to enhance the remote health management of scoliosis.

## 1. INTRODUCTION

Scoliosis is a complex 3D deformity of the spine characterised by an abnormal lateral curvature of the spine combined with the loss of normal sagittal curves ^1^ . It is also accompanied by a fixed vertebral and ribcage rotation component ^2–4^. This results in an externally observable torso distortion which affects normal posture ^5,6^. Scoliosis drastically diminishes quality of life in adulthood if left untreated ^7^, and in severe cases can cause back pain and negatively impact pulmonary and cardiac function ^2,8^. Scoliosis is the most common spine deformity ^9,10^, often presenting in childhood, and its prevalence varies depending on the aetiology, gender, age of onset, and geographical region ^10,11^.

Scoliosis is clinically diagnosed and monitored using two key metrics - the Cobb angle and the Angle of Torso Rotation (ATR). The Cobb angle is a measure of the lateral curvature, calculated from radiographic imaging in standing, and is the primary metric that is used for diagnosis and treatment decisions ^12^. The ATR is a measure of the vertebral rotation component and is non-radiographically inferred by quantifying the rib prominence in the posterior torso, typically measured when bending forward (Adam’s Forward Bend Test ^13^ depicted in Fig 1). It is the primary method of identifying possible scoliosis in children before confirmation with radiography and is used throughout the patient’s care trajectory to track deformity progression, the effect of various interventions and surgical success alongside the Cobb angle ^14^.

**Fig 1.**
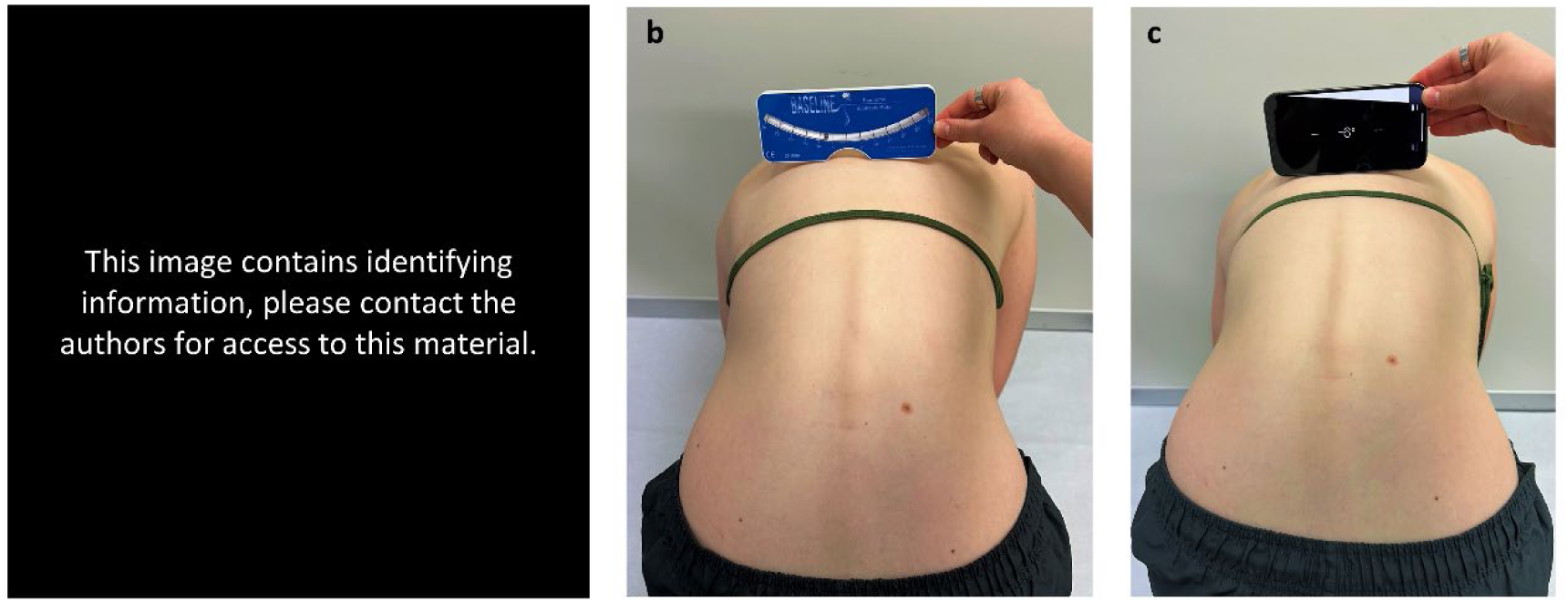
a) the Adam’s Forward Bend Test, b) combined with the use of the analog scoliometer. c) Sometimes, a smartphone inclinometer is used as a proxy for the analog scoliometer.

In the forward bending position, the ATR is measured using a simple inclinometer device called the scoliometer, developed by Bunnell for clinical use in 1948 ^15^. A measure of 7° or more is indicative of a scoliotic curve and a change of more than 5° is considered significant for a scoliosis patient ^15^. The reliability and accuracy of scoliometer measurements have been described controversially in the literature with the average error ranging from 5-8° with a variety of error sources ^14,16–21^. Commonly cited sources of error include variability in examiner experience and incorrect patient positioning ^16^. The most cited limitation of the device is the moving analog display and associated parallax error ^16^. In addition, there are some practical constraints. Firstly, the 30° scale limit is sometimes exceeded in patients with severe torso rotations. In these cases, clinicians will at times use the digital inclinometer on their smartphones as a proxy scoliometer, which is less accurate (see Fig 1) ^22^. Second, it is difficult to make accurate measurements when the patient’s torso cannot be positioned parallel to the ground (as sometimes observed in stiff post-op patients) as the functioning of the scoliometer (analog or smartphone based) relies on gravity and the orientation of the device relative to the ground. Lastly, there is some concern that the scoliometer does not span the full rib prominence in adults, larger males and/or high BMI patients.

Despite this, the concept and use of the scoliometer has remained relatively unchanged since its invention, with only minimal effort invested into its digitisation and automation. While the development of digital scoliometer devices ^23–26^ and smartphone apps ^18,22,27,28^ has reduced the measurement variabilities associated with the interpretation of scale-limited, parallax-prone analog readings, the clinical translation of such devices has been limited. Furthermore, remote measurement of the ATR is still not possible as even digital scoliometers and smartphones need to be positioned directly on the patient.

3D surface scanning (3DSS) is an imaging modality that uses structured light to produce 3D virtual surface topography information of a subject ^29^. It has garnered significant interest within the medical community in recent years because of its demonstrated simplicity, accuracy, and reliability in obtaining the 3D shape of a patient’s torso and extracting useful information from it ^29–35^. With the ever-improving quality of image capture, handheld 3DSS and photogrammetry devices are gaining popularity in remote health and telehealth applications because of their portability and ease of use ^29–32^.

Scoliosis progression can be effectively controlled when diagnosed early, consistently monitored by a spine specialist (in person clinic visits are essential for this) and treated in a timely manner ^10,36^. However, this convenience is not easily afforded to those living in remote or rural areas ^37–40^. Travel to a centrally located tertiary spine clinic places significant temporal, financial and emotional stress on patients and their families in addition to the costs incurred by the health service. These challenges lead to delayed diagnoses and limit conservative ‘early intervention’ treatment options, resulting in increased rates of avoidable surgical correction. While the need for competent remote health management for rural and remote communities has always been evident ^41^, the Covid-19 pandemic has provided much needed impetus for clinics to adopt the use of “digital twins” in telehealth protocols ^42^. With recent advances in digital imaging technology and computing power, there is now a renewed interest in using virtual data for the remote assessment of scoliosis, particularly in children.

To our knowledge, no virtual scoliometers have been explored to date. This paper describes the first development and clinical validation of a digital twin for the analog scoliometer that uses patient-specific 3DSS data. We investigate the reliability, accuracy, sensitivity and specificity of the tool, and its potential for future clinical use. The use of a generative design algorithm that allows parametric user control is an effective way to introduce automation into the measurement process and streamline the analysis of patient-specific virtual data, potentially increasing the accuracy of the measurements and decreasing the variance between users. The development of such a digital twin is expected to have significant clinical value in rapidly emerging digital health management technologies for remote spine deformity care.

## 2. METHODS

Adolescent Idiopathic Scoliosis (AIS) is the most common type of scoliosis ^43^ affecting approximately 4% of the adolescent population ^44^ and will be a profuse patient group for uptake of this new digital tool. The digital scoliometer tool was developed using a graphical coding environment to replicate the physical measurement process of ATR with the analog scoliometer, using 3D virtual models of AIS patients and non-scoliotic, healthy developing adolescents. It is packaged into a simple graphical user interface for user-friendliness. Details of the recruited participants, tool development and verification/validation of the tool are detailed below.

### 2.1. Datasets

Three cohorts were considered in the current study - a group of non-scoliotic, healthy developing adolescents (control), a group of AIS patients selected from a historical database of AIS patients treated at the Queensland Children’s Hospital Spine Clinic (QCHSC), and a group of operatively treated AIS patients also selected from this same historical database, for whom both pre- and post-operative clinical data was available. Participant demographics are mapped out in Table 1 and the inclusion criteria for each dataset are listed below.

**Table 1.** Demographics of healthy participants and AIS patients that participated the study.

| Demographics | Dataset 1<br>(Healthy) | Dataset 2<br>(AIS first presentation) | Dataset 3<br>(AIS surgical) |  |
| --- | --- | --- | --- | --- |
|  | (n=40) | (n=40) | Pre-op<br>(n=35) | Post-op<br>(n=35) |
| **Gender** |  |  |  |  |
| Male | n=18 | n=6 | n=4 |  |
| Female | n=22 | n=34 | n=31 |  |
| **Age (years)** |  |  |  |  |
| Mean ± SD | 13.6 ± 1.9 | 13.8 ± 1.9 | 14.9 ± 1.6 | 15.1 ± 1.6 |
| Min | 11 | 9.6 | 11 | 11.2 |
| Max | 18 | 17.28 | 17 | 17.3 |
| **BMI (kg/m<sup>2</sup>)** |  |  |  |  |
| Mean ± SD | 20 ± 3 | 19.8 ± 3.6 | 19.9 ± 3.4 | 19.2 ± 3.2 |
| Min | 14.8 | 13.5 | 14.9 | 14.8 |
| Max | 25.5 | 30.3 | 30.7 | 29.2 |
| **ATR major curve (°)\*** |  |  |  |  |
| Mean ± SD | 3.5 ± 2.6 | 7.8 ± 2.5 | 21 ± 6.3 | 10 ± 4.4 |
| Min | 0 | 2 | 10 | 0 |
| Max | 13 | 11 | 38 | 21 |
| *\*ATR measured using standard clinical method with the analog scoliometer.* |  |  |  |  |
| **Cobb angle (°)** |  |  |  |  |
| Mean ± SD |  | 29.7 ± 11.1 | 67 ± 12.8 | 28 ± 9.2 |
| Min |  | 10 | 48 | 13 |
| Max |  | 58 | 105 | 56 |
| **Lenke type** |  |  |  |  |
| 1 |  | n=26 | n=21 |  |
| 2 |  | n=1 | n=2 |  |
| 3 |  | n=5 | n=6 |  |
| 4 |  | n=0 | n=1 |  |
| 5 |  | n=8 | n=4 |  |

#### Dataset 1: Healthy non-scoliotic participants

40 healthy developing adolescents between the ages 10 to 18 years, with no history of diagnosed spine deformity or disorders, lower limb conditions or prior spine surgery were recruited as controls. Eligible volunteer participants were scanned under standard scanning protocols developed within the research group (described in the next section).

#### Dataset 2: AIS patients at first clinical presentation

This dataset was a subset from the existing historical 3DSS database of AIS patients treated at the QCHSC. A subset of scans from the database were selected at the first timepoint when the patients initially presented at the spine clinic. Scans were only chosen if the clinically measured ATR was 10° or less, if the patient was subsequently diagnosed with AIS, and received treatment for the same.

#### Dataset 3: AIS patients treated surgically

A subset of 35 patients scanned 1 week pre- and at 2 months post-operatively were chosen from the existing historical 3DSS database of AIS patients treated at the QCHSC.

### 2.2. Scan protocol

The existing historical database of 3DSS data for AIS patients followed an optimised scan capture protocol. Before each scan, the patient’s height, weight, and date of birth were recorded. Patients were scanned in the Adam’s forward flexion position at 22 frames per second using the Artec Leo scanner (Artec Group Inc., Luxembourg). Males were scanned shirtless while females were asked to change into crop tops provided to them in clinic by the researcher. All patients were asked to stand on a wooden calibration board designed and fabricated in-house, which provided a reliable co-ordinate frame for later alignment of the digital scan data. A rectangular box support was also provided for stability, which the patient could ‘rest’ their hands on to prevent them from swaying during the scan (as shown in Fig 1a). The scan path was kept consistent, following a 360° path, with the feet and board captured first, followed by the legs, torso and finally the shoulders. Each scan roughly takes a minute.

We replicated this protocol for the participant group of healthy developing adolescents, to ensure consistency between scanning and the resultant 3D virtual models.

### 2.3. Scan processing

The raw scans were stitched in the associated software Artec Professional (Version 17), and the fused, watertight 3D models were exported in a binary stereolithography (.stl) format to be further processed in Geomagic Wrap 2021 (Oqton, USA) using a standardised workflow developed in-house. Here the models were aligned to the world coordinate system using the co-ordinate frame that was captured on the wooden calibration board on which the patients stood to be scanned. The models were then cropped to only include the torso (head and limbs were removed) and any unwanted items captured in the background were deleted. Next, the model was globally smoothed. Excessive smoothing and noise reduction can result in loss of detail, so this operation was only performed once per model. The inbuilt “Mesh Doctor” tool was used to repair errors (such as non-manifold edges, self-intersections, highly creased edges, spikes, small components and small holes) if any, in the polygon surface mesh. The mesh was then conservatively decimated to reduce the file size. In preparation for further processing with the digital tool (detailed below), the anterior torso was cropped to a flat surface, leaving only the posterior torso from shoulder to just below the left/right posterior superior iliac spines (PSIS), as this is the region of primary interest when measuring ATR using a scoliometer. An example of a fully processed, calibrated and deidentified 3D model is shown in Fig 2.

**Fig 2.**
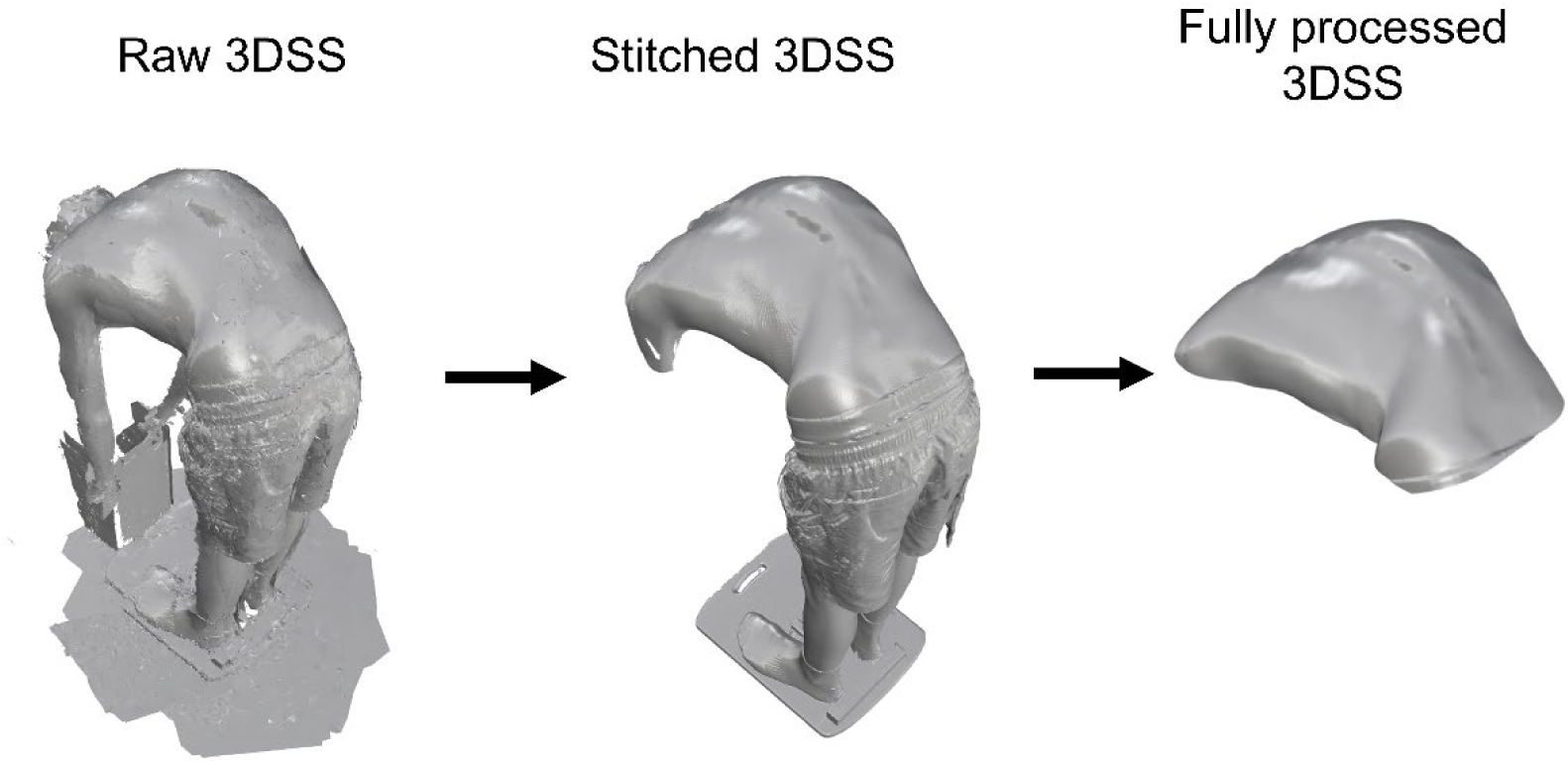
3DSS scan processing of participant in the forward bending position whilst standing on the calibration board and using the anti-sway support.

### 2.4. Algorithm development

A virtual digital scoliometer tool was developed to measure ATR from 3DSS data using the 3D modelling and graphical programming software Rhino3D-Grasshopper (Robert McNeel and associates, USA). The tool is of a generative and parametric design, allowing a user to make semi-automated measurements in a controlled and streamlined manner, while at the same time manipulating the orientation, perspective view and position of the 3D model in the Rhino workspace ^30^. User inputs to the algorithm can be made through the graphical user interface (GUI) on the Grasshopper canvas. The user must follow three successive, simple steps detailed below (and displayed in Fig 3) to calculate ATR values:

Step 1 – Import the mesh model into Rhino by dragging the .stl file into the workspace. Place a minimum of 5 points (there is no maximum) on the spine, from which the algorithm will automatically estimate the spine curve (scoliosis).
Step 2 – Use the number sliders to define a region of interest (the rib prominence) on the estimated spine curve from which to measure the ATR.
Step 3 – Indicate the lateral torso limits across which the ATR will be measured, using the number sliders.

**Fig 3.**
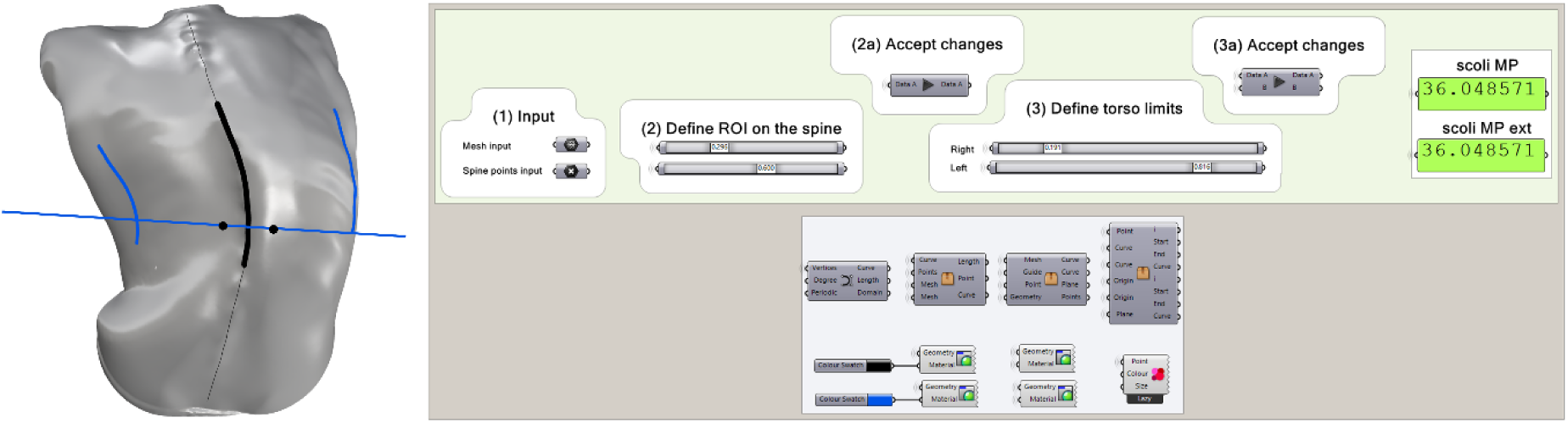
3DSS torso model of a scoliosis participant in the ‘forward bend position’ in the Rhino viewport showing an example measurement (left) with the corresponding GUI on the Grasshopper canvas (right). The output box (green) displays the calculated ATR. Note: The user need not interact with the cluster of grey boxes in the lower half of the Grasshopper canvas – only the slide bars in the upper half.

The steps detailed above take approximately 30 seconds to complete per scan. An example measurement on the Rhino viewport and Grasshopper user interface is shown in Fig 3. ‘Scoli MP’ shows the calculated ATR value when the virtual scoliometer has the same dimensions as the analog device. A multiplanar measurement is made from the curved trajectory of the patient’s spine curve. ‘Scoli MP Ext’ works similarly but has an *extended device length* to span the full torso of the patient.

The logic of the algorithm is pictorially depicted in Fig 4. The user is required to place a minimum of five points on the input mesh to indicate the approximate spine trajectory. A curve is created from this and divided into 1 mm segments. A search function is then deployed where the algorithm finds the closest point on the mesh to each division point on the curve. A new high-resolution curve is finally created with this increased number of input points. This curve forms the base on which the user defines the region of interest for ATR measurement.

**Fig 4.**
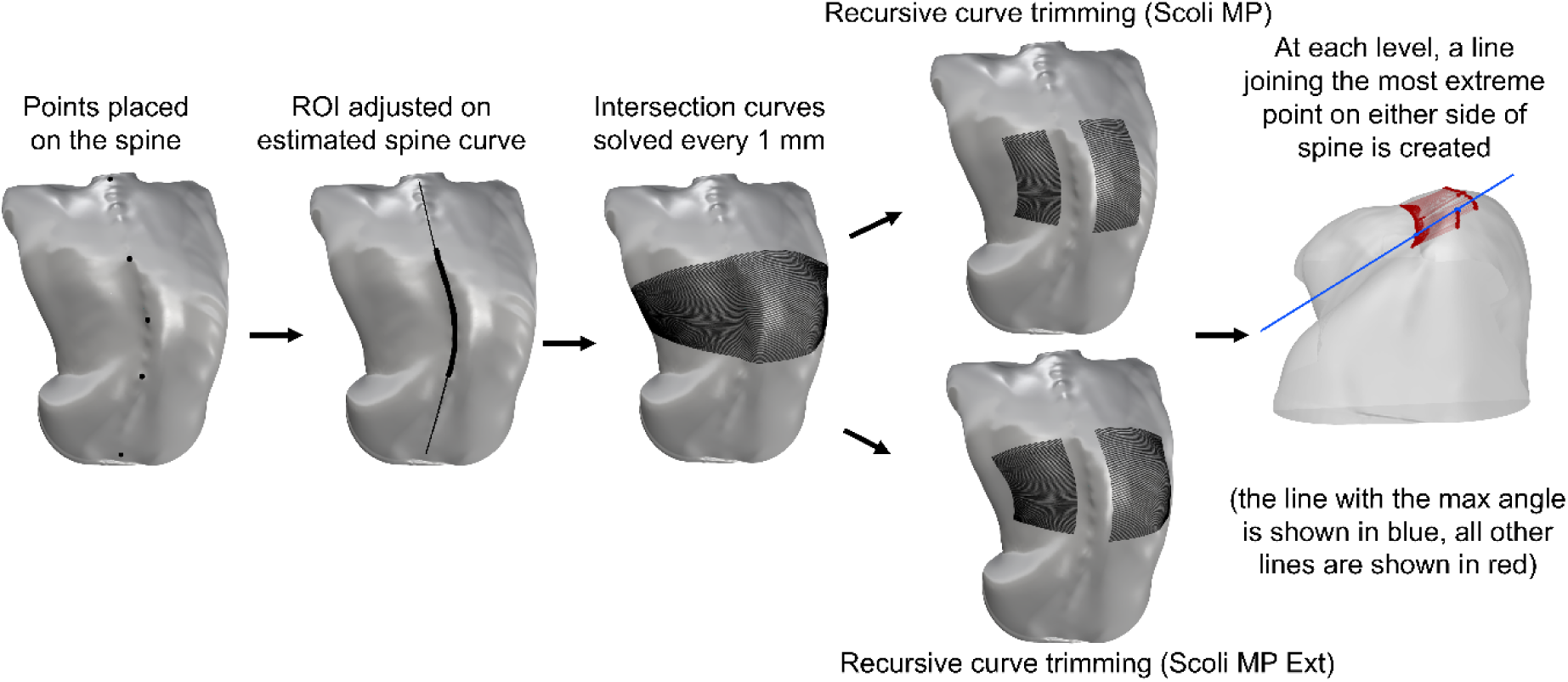
Algorithm logic of the digital twin described step by step (the model showing the last step of the process is made partially transparent for viewing clarity of the lines). The generative design of the workflow allows for any calibrated model to be measured in this manner.

Once the ROI is defined, the algorithm creates planes perpendicular to the curve trajectory at each input point. A Mesh-Plane intersection is solved for every plane, which creates an intersection curve every 1 mm along the ROI. The curve is then recursively trimmed to create sub-curves according to the length of the analog scoliometer (‘Scoli MP’) and according to the user input of the torso limits (‘Scoli MP Ext’). In both versions, an allowance for the spine is provided, measuring the same length as the one found on the analog scoliometer.

On each intersection curve, on both sides of the spine, the most extreme point is identified. A line is drawn at each level joining the two extreme points. The angle of each line is measured, and the line with the maximum angle is extracted. This angle is the ATR measurement displayed in the output box.

A semi-automated scoliometer tool to digitally measure the ATR from a virtual 3DSS model was thus successfully developed.

### 2.5. Measurement and analysis protocol

Fig 5 gives an overview of the different datasets used for the analyses listed below.

**Fig 5.**
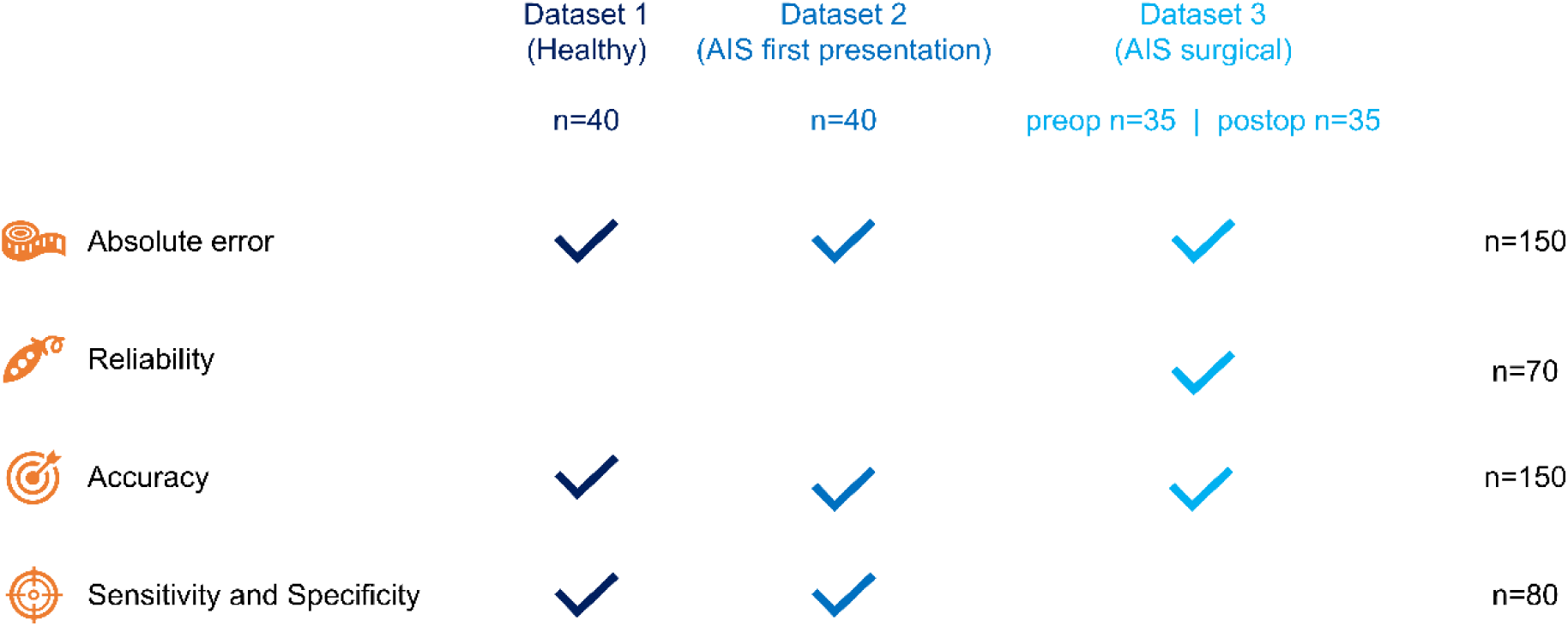
Chart detailing dataset usage for the four analyses.

#### Absolute Error

This analysis aimed to investigate what is gained by increasing the length of the virtual device. The absolute error was calculated between ‘Scoli MP’ and ‘Scoli MP Ext’ to determine whether increasing the length of the digital scoliometer to span the entire torso produced significantly different values to those obtained from the standard device length.

#### Reliability

This analysis aimed to investigate measurement repeatability when ATR was calculated by a single user and by multiple users. For inter-user reliability assessment, three users were each asked to record ATR measurements once. One user also completed the full set of measurements two additional times for intra-user reliability assessment. Measurements were completed over the span of a few weeks to mitigate any memory bias. Inter-Class Correlation was then calculated form these measurements to assess if the tool produces repeatable measurements regardless of the user.

#### Accuracy

This analysis aimed to compare how close the digital measurements were to the clinically reported analog measurements. For the successful clinical uptake of such a virtual digital scoliometer tool, it is important to show that the measurements are comparable to those obtained with current clinical practices. For the scoliosis cohorts, the analog measurement was made by the QCH clinical staff during outpatient spine clinic appointments on the same day as their 3DSS scan. For the healthy non-scoliotic cohort, the analog measurement was made at the time of their 3DSS by a highly experienced QCHSC physiotherapist specialising in AIS.

A correlation matrix was calculated to obtain the relationship between the digital and analog scoliometer measurements. To further assess agreement in the values themselves, a Bland-Altman analysis was carried out by denoting the analog ATR values as the ‘reference’.

#### Sensitivity and Specificity

This analysis aimed to test the robustness of the algorithm when measuring ATRs of low magnitude. For Dataset 1, if the analog measurement was ≤ 7°, it was considered a true negative, and if measurement was >7° it was considered a true positive. This allowed for the possibility that a participant presenting as a healthy non-scoliotic participant may have undiagnosed scoliosis. For Dataset 2, if a measured ATR > 7°, it was considered a true positive.

### 2.6. Statistical analysis

All statistical analysis was performed on R statistical computing software (R Foundation for Statistical Computing, Vienna, Austria) ^45^. The error bars describe standard deviation.

Intra- and inter-user reliabilities were calculated using a two-way, random effects, absolute agreements, single rater Inter-Class Correlation (ICC) according to the McGraw and Wong classification. Sample size was calculated with a minimum accepted ICC reliability of 0.5 and an expected reliability of 0.8 ^46,47^. Significance level was set at 0.05 (two-tailed) at a power of 80%. ICC values of 0.5 and above indicate that the method is reliable. In detail, poor reliability is indicated by values less than 0.5, moderate reliability is indicated by values between 0.5 and 0.75, good reliability is indicated by values between 0.75 and 0.9, and excellent reliability is indicated by values greater than 0.90.

Spearman correlation coefficient was used to create the correlation matrix to account for non-linearity in the data. Significance levels were denoted as * (p < .05), ** (p < .01) and *** (p < .001).

Limits of Agreement (LoA) for the Bland Altman analysis were calculated at a significance level of 0.95 as is standard.

Sensitivity, specificity, positive predictive value (PPV), and negative predictive value (NPV) were calculated using standard formulae.

### 2.7. Ethics statement

Hospital and University Human Research Ethics Committee (HREC) approvals were obtained from QCH HREC (LNR/21/QCHQ/75249) and the Queensland University of Technology HREC (Approval number: 4856 – HE44) titled “Spine Deformity Management Clinical Data Collection Project”. Approval to publish de-identified group data analyses by the QCH HREC for the “Development of non-invasive monitoring tools for Adolescent Idiopathic Scoliosis, using 3D scanning/photography at the Queensland Children’s Hospital” was also provided. Informed consent was obtained from all participants and their legal guardians.

The methods described in this paper are in accordance with the relevant guidelines and regulations put forth by QCH and QUT HREC and the Declaration of Helsinki.

## 3. RESULTS

### 3.1. Absolute error

The mean absolute error between ‘Scoli MP’ and ‘Scoli MP Ext’ measurements was 0.023 ± 0.13° with errors ranging from a minimum of 0° to a maximum of 1.36°.

### 3.2. Reliability

ICC values for inter and intra-user reliability are graphically represented in Fig 6. Inter-user reliability was excellent for ‘Scoli MP’ (0.978) and ‘Scoli MP Ext’ (0.981). The intra-user reliability was also excellent for ‘Scoli MP’ (0.991) and ‘Scoli MP Ext’ (0.991).

**Fig 6.**
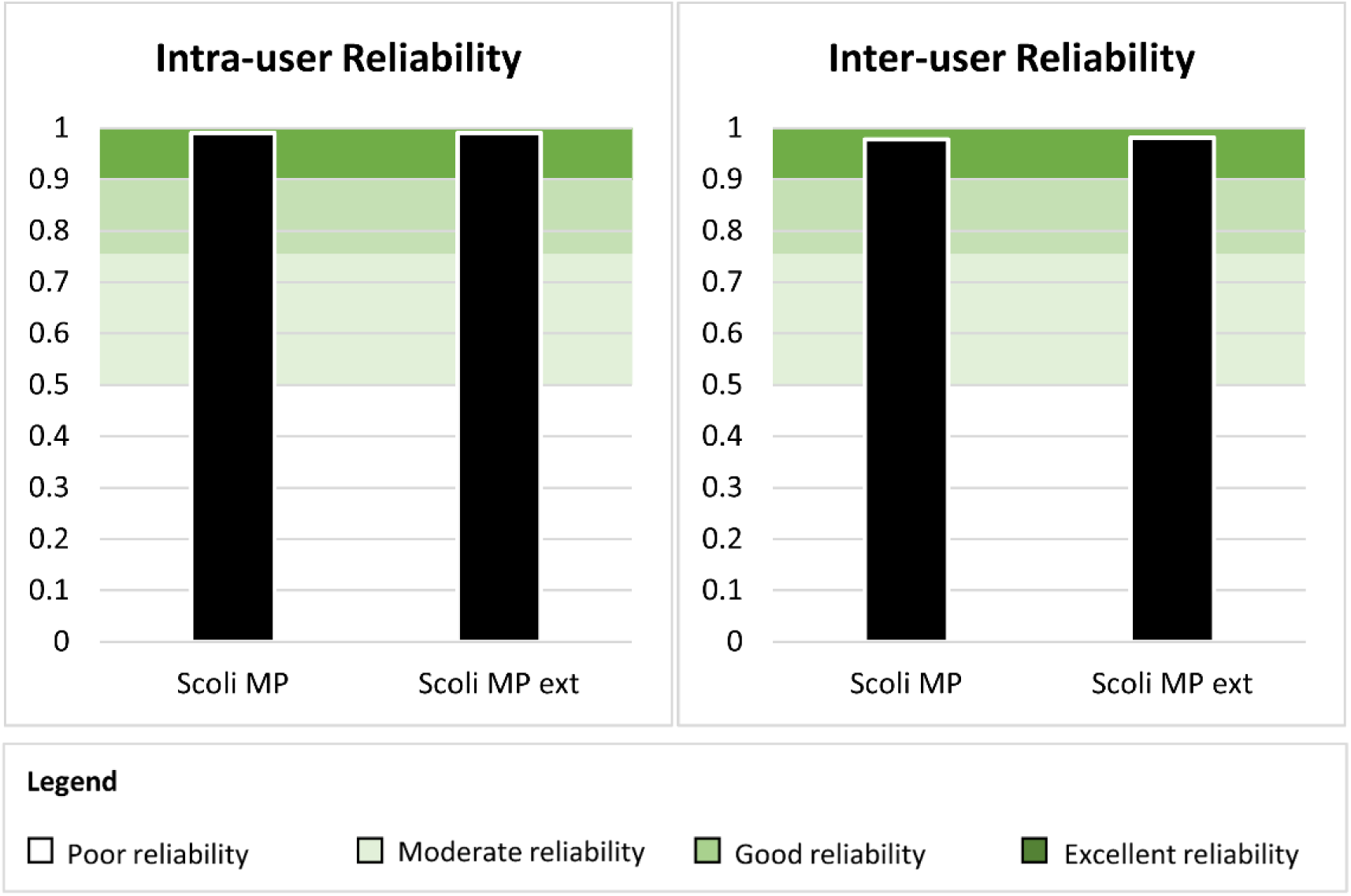
Graph showing ICC values for inter- and intra-user reliability.

### 3.3. Accuracy

A correlation matrix with Spearman’s correlation coefficient for the digital values versus the clinical analog values is shown in Fig 7. Both ‘Scoli MP’ and ‘Scoli MP Ext’ showed a high positive statistically significant correlation with the clinically measured ATR values with coefficients of 0.897 and 0.896 respectively.

**Fig 7.**
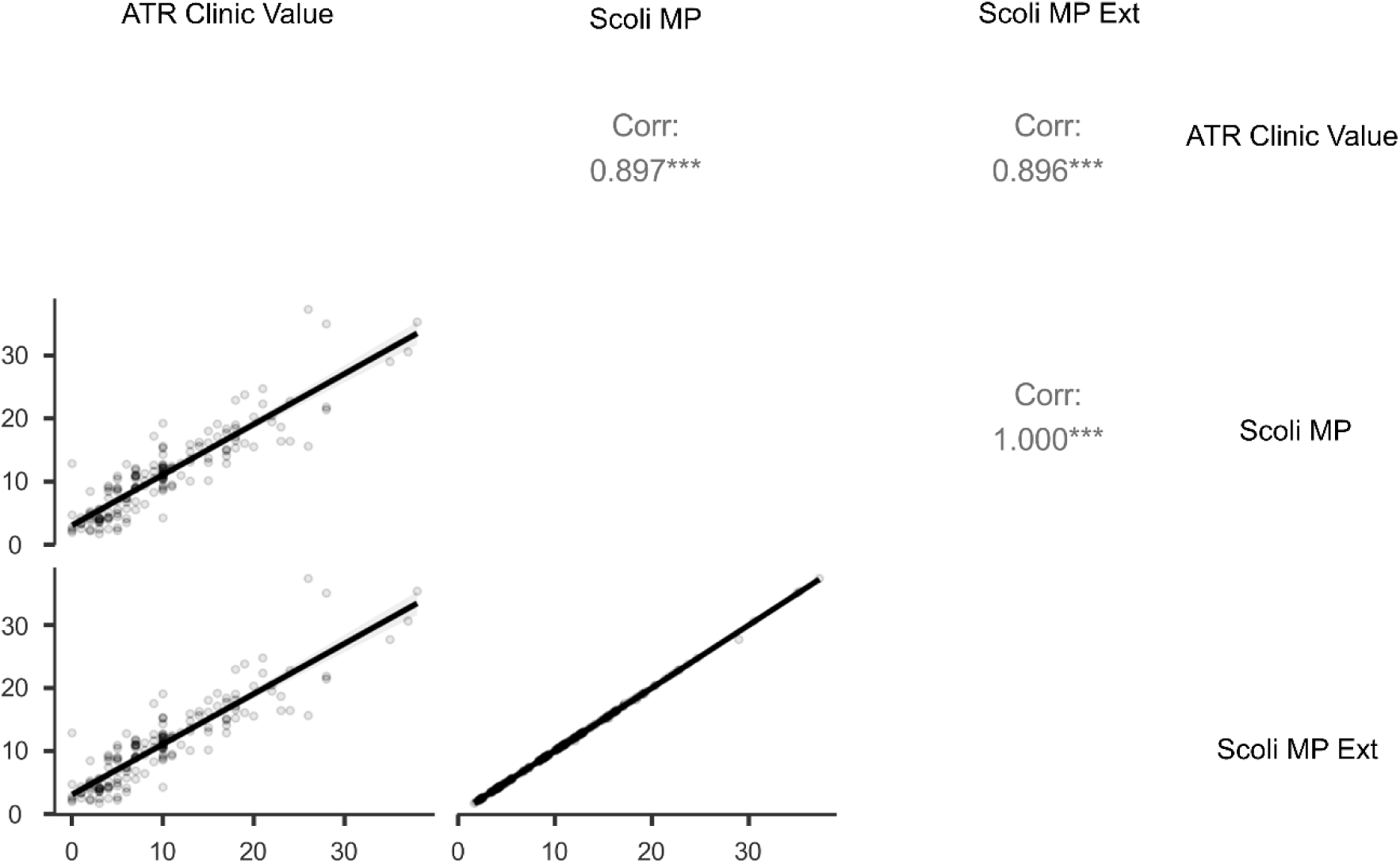
Spearman correlation matrix comparing ‘Scoli MP’ and ‘Scoli MP Ext’ with clinic ATR values (p<0.001 is shown as ***)

Bland Altman analysis was conducted to further investigate the agreement between the digital values and the analog ATR values (Fig 8). On average, ‘Scoli MP’ measures 0.98° higher than the corresponding analog ATR values and ‘Scoli MP Ext’ measures 0.96° higher than the corresponding analog ATR values. Both plots reveal minimal proportional bias that approaches zero around a mean ATR of 19°. 92.7% of measurements were within the LoA in both plots.

**Fig 8.**
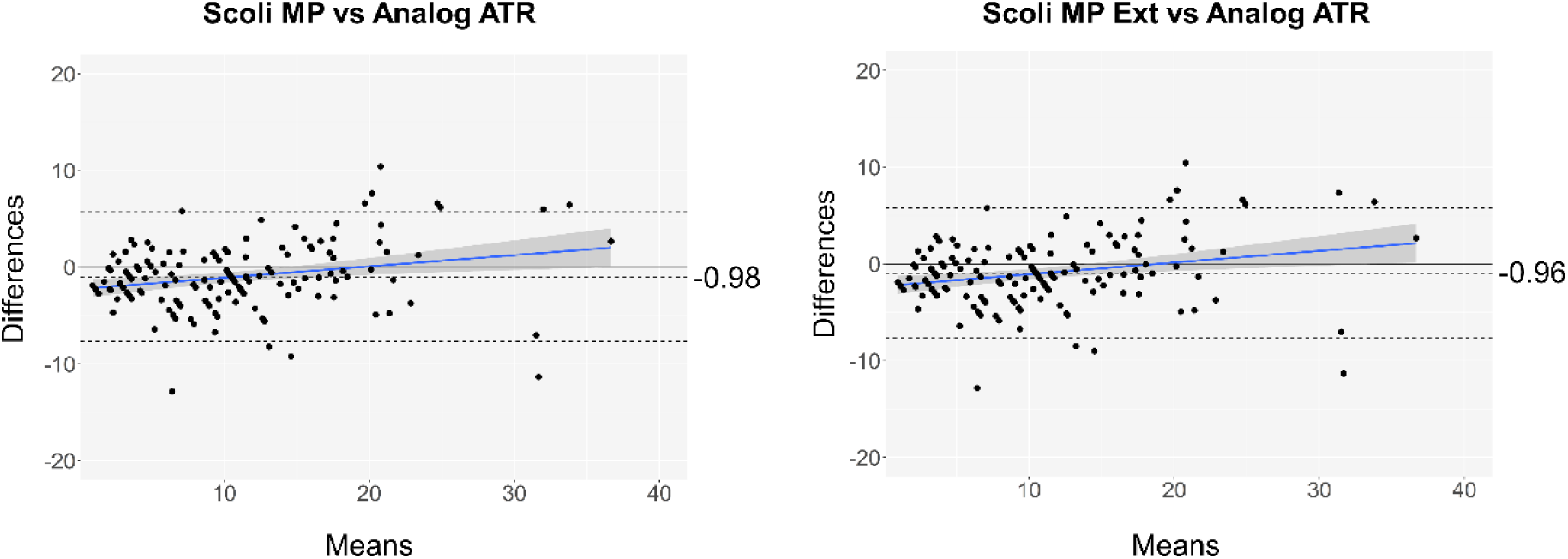
Bland-Altman plots for ‘Scoli MP’ and ‘Scoli MP Ext’ vs analog scoliometer values (reference). Average bias is -0.98 for ‘Scoli MP’ and -0.96 for ‘Scoli MP Ext’. LoA are calculated at 0.95 confidence level.

### 3.4. Sensitivity and specificity

The algorithm showed a sensitivity of 95.83%, a specificity of 76.76%, a PPV of 63.89% and an NPV of 97.72%.

## 4. DISCUSSION

The Cobb angle, measured on a standing radiograph, is the primary metric used for scoliosis diagnosis, monitoring of scoliosis progression, conservative or surgical treatment choices and finally to measure the effect (if any) of any treatment that was prescribed ^48^. However, it does not fully describe the severity of the AIS deformity in three dimensions. While there has been significant effort invested into determining multi-planar deformity correlation in the progressively deforming AIS spine, these correlations are weak and the various deformity elements must still be analysed separately ^16^. Quantification of axial rotational deformity and the resulting ATR therefore still holds considerable importance in the holistic assessment of scoliosis manifestation and treatment in a patient with scoliosis.

Over many years, there have been several attempts at developing a wide range of both radiographic as well as non-radiographic methods for measuring the intrinsic vertebral rotation that is a key feature of scoliosis ^49^. Using radiography, the popular methods are the Nash-Moe method ^50^, the Perdriolle method ^51^, the Stokes method ^52^, the Raimondi method ^53^, and the Rib Vertebra Angle Difference method (for infants) ^54^. Most notable non-radiographic and non-invasive methods include the Adam’s forward bend test (with and without the scoliometer) ^13^, Moiré topography ^55^, spinal ultrasound ^56^, depth sensors ^57,58^, infrared thermography ^59^ and techniques based on 3D surface topography ^60–64^. It must be noted that radiographic methods are predominantly used with the patient in the standing position and non-radiographic methods are predominantly used with the patient in the forward bending position which more clearly displays the ATR produced by scoliosis.

And yet, the most widely used tool to measure axial rotation in scoliosis or for scoliosis screening continues to be the analog scoliometer or a smartphone mimicry of it. While the scoliometer was first adopted to reduce radiation exposure to young AIS patients, it is now used ubiquitously in clinics as a simple and inexpensive method of diagnosing and monitoring scoliotic rotational deformity progression alongside traditional radiographic imaging ^15^. Despite the widespread acknowledgement of its measurement variability ^14,16,65^, it remains the gold standard method for ATR measurement in scoliosis centres across the world.

The work presented here is the first development of a digital twin framework for the analog scoliometer and a full validation of its reliability, accuracy, sensitivity, and specificity. With simple user inputs and a patient-specific virtual 3D model, the ATR can be measured digitally in seconds. The tool was developed on the Rhino3D-Grasshopper commercial software which is affordable and easily usable in a generic computer. Rhino3D was developed primarily for use in the architecture field to enable the parametric design of buildings but is now gaining popularity in other fields due to its ability to create generative algorithms ^30,66^.

The digital tool was tested first for reliability and found to produce highly repeatable measurements both for a single user and multiple users. In addition, anecdotal feedback from one of the three users, a Training Spine Fellow and Orthopaedic Surgeon at the QCHSC, indicated excellent user friendliness of the tool with respect to ease of input and low measurement time per scan.

The ATR measurements produced on the digital tool showed a strong positive correlation with the clinically measured scoliometer ATR values, indicating that it would be a good digital alternative to the current ‘gold standard’ analog measures. In addition, the digital ATR values were shown to have excellent agreement (92.7%) with the analog ATR values (having 95% points within LoA indicates that the two methods being tested are interchangeable ^67^). The average bias of less than 1° between the digital and analog methods is negligible.

The digital tool also shows evidence of satisfactory performance as a screening tool with a combined sensitivity and specificity value of 1.72 (a value of 1.5 generally indicates that the test method is useful ^68^). However, in this study, we have taken an equal number of healthy non-scoliotic controls and AIS patients (50% prevalence), resulting in some spectrum bias. Since sensitivity, specificity, PPV and NPV results are sensitive to prevalence, we expect that in clinical practice the PPV will increase and the NPV will decrease to reflect the much lower prevalence of scoliosis in the general population ^69^.

In Dataset 1 (healthy non-scoliotic participants), three participants were found to have an analog ATR measurement higher than 7° indicating possible scoliosis. As per our ethical approvals, they were referred for assessment by their General Medical Practitioner. For the purpose of assessing sensitivity and specificity, they were classified as ‘true positives’ despite being part of the healthy non-scoliotic dataset.

In addition to the validation presented in this paper, it is worth noting that the virtual digital tool is not constrained by gravity as is the case for the analog scoliometer. It can measure ATR regardless of the positioning of the torso relative to the ground, enabling the quantification of torso distortion with the patient in a more upright standing position if required. This not only allows for error-free ATR measurement of stiff post-operative patients, or those with limited Hamstring muscle length, who find it difficult to correctly position their bodies for the Adam’s forward bend test, but also enable ATR measurements in the standing position should this be of clinical relevance in determining cosmetic deformity. Further, the results show that increasing the length of the digital tool to span the full breadth of the patient’s torso did not significantly alter the measurements. While it is a useful feature of the tool, its practical relevance is not evident, as the original length of the analog scoliometer device was not shown to be a disadvantage.

## 5. CONCLUSION

In conclusion, this work has established a streamlined digital twin for the analog scoliometer that uses a semi-automated algorithm to measure ATR from patient-specific 3DSS data. The digital twin mimics the process of manually following the spine curve with an analog scoliometer to find the position of maximum asymmetry in a scoliotic torso. The tool is fast, reliable, accurate, sensitive, specific, not limited by gravity, can measure angles higher than 30°, and is not affected by parallax errors. While it was necessary to validate the tool on a high accuracy 3DSS model for the purpose of the study, it can potentially be used to measure the ATR on a 3D model of a patient obtained from any source. Future work would involve testing the agility of the tool with lower quality inputs like those from smartphone photogrammetry or depth sensors. This is expected to be hugely beneficial for implementation in telehealth initiatives for remote diagnosis and management of scoliosis and other spine deformities.

## DATA AVAILABILITY STATEMENT

The raw measurement data and statistical analysis made on the algorithm will be made fully available to the public via QUT’s public repository Research Data Finder (RDF) after the paper has been published. Through the RDF, noninstitutional researchers may be granted access to this data after making a request to myself as first author (Dr Sinduja Suresh) and owner of the RDF entry.

The Rhino-Grasshopper workflow developed in this project is not yet available for public dissemination because we are currently exploring opportunities with local hospitals to include it in a smart device application. However, the workflow is built on commercially available software, and we have described the functionality in detail. We believe that someone with sufficient experience in Grasshopper could replicate this functionality described in the Methods section.

The original 3DSS datasets cannot be shared publicly as it may compromise the privacy and confidentiality of our participants.

## ACKNOWLEDGEMENTS

We thank Miss Rachel Chalmers, Dr Caroline Grant, and Miss Daniela Blomberger for scanning the majority of the patients listed in our 3DSS AIS database and all the Work Integrated Learning (WIL) students for performing a portion of the mesh post-processing. The authors are also grateful for the discussions and input from the rest of the Orthopaedic Advisory Board and BSRG team.

## AUTHOR CONTRIBUTIONS

**SS** – conceptualisation, data curation, formal analysis, investigation, methodology, project administration, software, visualisation, writing (original draft preparation, editing)

**AS** – data curation, formal analysis, investigation, methodology, visualisation, writing (review, editing)

**AA** – data curation, writing (review)

**MI** – project administration, resources, supervision, writing (review, editing)

**RL** – supervision, validation, writing (review)

**GA** – supervision, validation, writing (review)

**JPL** – conceptualisation, project administration, resources, supervision, writing (review, editing)

